# Integrating Telerehabilitation into the Prehabilitation and Rehabilitation Pathway in Colorectal Cancer: A Protocol for a Randomized Controlled Trial

**DOI:** 10.1101/2025.09.18.25336078

**Authors:** José Manuel Burgos-Bragado, Carolina Jiménez-Sánchez, Natalia Brandín-de la Cruz, Beatriz Carpallo-Porcar, Lilian Le-Roux, Juan Luis Blas-Laina, Jorge Alamillo-Salas, Paola Gracia-Gimeno, Sandra Calvo

## Abstract

**Introduction:** Colorectal cancer (CRC) is a leading global malignancy, and surgery is frequently followed by complications, functional decline, and reduced quality of life. Multimodal prehabilitation and rehabilitation can improve physical recovery and psychosocial outcomes, but uptake is often limited by logistical and mobility barriers. Asynchronous telerehabilitation provides a flexible, patient-centred and scalable solution, yet its effectiveness across the perioperative CRC pathway has not been rigorously evaluated. This trial will test a multimodal asynchronous programme, delivered in prehabilitation and postoperative phases, against a standard leaflet-based approach.

**Methods:** This single-blind, parallel-group randomized controlled trial comparing an asynchronous, multimodal telerehabilitation program with a leaflet-based standard program in adults scheduled for elective CRC resection. Fifty-six participants will be randomized 1:1 to telerehabilitation (HEFORA platform) or control. The intervention spans a 2-week prehabilitation phase and a 4-week postoperative phase. Assessments occur at five time points: baseline (pre-prehabilitation), post-prehabilitation (pre-surgery), post-surgery (pre-rehabilitation), post-rehabilitation, and 3-month follow-up.

**Outcomes:** The primary outcome is functional capacity (Six-Minute Walk Test distance). Secondary outcomes include muscle strength, body composition, pulmonary function, physical activity, sleep quality, psychosocial measures, health-related quality of life, treatment expectancy, usability, satisfaction, and adherence. Analyses will follow the intention-to-treat principle with appropriate models for repeated measures and sensitivity analyses for missing data and adherence.

**Conclusions:** This study will provide essential evidence on the role of asynchronous telerehabilitation in perioperative colorectal cancer management. Positive results could inform clinical guidelines, promote wider adoption of digital rehabilitation strategies, and drive a more accessible, patient-centered, and cost-effective approach to oncologic recovery.

**Trial registration:** ClinicalTrials.gov identifier: NCT06593678

**Ethics approval:** Aragón Clinical Research Ethics Committee (PI23/557).

## 1. Introduction

Colorectal cancer (CRC) is a malignant neoplasm arising from the inner lining of the colon or rectum, most commonly as adenocarcinomas. It follows the well-established “adenoma-carcinoma sequence”, in which adenomatous polyps progress to invasive carcinoma through the accumulation of driver mutations in genes such as APC, KRAS, TP53 mutations and epigenetic alterations[1]. Clinically, tumor growth may lead to luminal obstruction, bleeding, and metastasis spread, with symptomatology varying by tumor location due to differences in vascular and lymphatic drainage pathways (e.g., ascending vs. transverse vs. descending vs. sigmoid colon vs. rectum) [2]. Common clinical presentations include rectal bleeding, changes in bowel habits, abdominal pain, anemia, and unexplained weight loss, though early-stage disease is often asymptomatic, contributing to diagnostic delays when symptoms are misattributed to benign conditions, thus reinforcing the critical importance of screening [3]. Indeed, early detection through screening modalities such as colonoscopy or fecal testing can identify and remove pre-cancerous polyps or detect cancers at a treatable stage, substantially improving outcomes [4].

CRC represents a major health challenge due to its high incidence, substantial mortality rate, and the physical, psychological, and economic burden it imposes on patients and healthcare systems. According to the World Health Organization (WHO), CRC is the third most frequently diagnosed cancer and the second leading cause of cancer-related death worldwide, accounting for approximately 10% of all cancer cases and 9.6% of cancer deaths annually [5]. Globally, CRC affects more than 1.9 million individuals per year, with over 903,859 deaths annually [6]. In low- and middle-income countries, incidence rates are rising, largely driven by dietary westernization, reduced fiber intake, increasing consumption of highly processed foods, and sedentary lifestyle [7]. In high-income regions such as Europe and North America, organized screening programs have improved early detection, but the ageing population and persistent risk factors (unhealthy dietary patterns, alcohol consumption, obesity, and physical inactivity) sustain a disease burden [8]. The incidence increases significantly after the age of 50, and men are slightly more frequently affected than women [9].

Risk factors for CRC include both modifiable and non-modifiable determinants. Established modifiable factors comprise smoking, alcohol consumption, sedentary behaviour, obesity, low dietary fibre intake, and high consumption of red and processed meats [10]. Non-modifiable factors include age, family history and hereditary syndromes such as Lynch syndrome and familial adenomatous polyposis [11].

The management of CRC requires a comprehensive, multidisciplinary approach encompassing surgical resection, chemotherapy, radiotherapy, and, when indicated, immunotherapy with immune checkpoint inhibitors. Current guidelines emphasize multidisciplinary team (MDT) involvement, especially through tumor board conferences, to optimize treatment decisions based on tumor characteristics and patient condition [12].

Treatment decisions are individualized based on tumor staging, molecular and genetic profiling, and patient-specific clinical factors [12]. Surgical resection, often accompanied by lymphadenectomy, remains the cornerstone of curative treatment and may involve procedures such as segmental colectomy or low anterior resection. In the rectal cancer, total mesorectal excision (TME) is the standard surgical technique, improving oncologic outcomes and sphincter preservation [13].

However, postoperative recovery is often impaired by pain, fatigue, sarcopenia, and emotional distress, all of which contribute to reduced functional capacity, particularly in frail or elderly patients, undermining treatment compliance and autonomy [14]. Despite advances in early detection, surgical techniques, and oncologic therapies, CRC surgery remains associated with significant morbidity and long-term functional decline, particularly in these vulnerable populations. These postoperative challenges not only delay convalescence and increase healthcare utilization but also affect overall quality of life (QoL) [15].

Considering these challenges, the implementation of structured rehabilitation programs, both in the preoperative phase (prehabilitation) and postoperatively, is crucial to optimize functional recovery, reduce postoperative complications, reduce healthcare costs, and enhance long-term patient outcomes [16,17].

Exercise and rehabilitation are recognized as core components of comprehensive cancer care, as endorsed by international clinical guidelines [18,19]. Multimodal prehabilitation, which combines structured physical exercise, nutritional optimization, and therapeutic education, has demonstrated significant benefits in improving preoperative functional capacity, reducing postoperative morbidity, and accelerating recovery trajectories in patients undergoing CRC surgery [20–22]. Likewise, postoperative exercise-based interventions play a key role in restoring physical function, facilitating recovery, and enhancing overall QoL [23].

These interventions show enhanced effectiveness when integrated into Enhanced Recovery After Surgery (ERAS) protocols, which are standardized, evidence-based perioperative care pathways aimed at minimizing surgical stress, maintaining physiological function, and expediting recovery. ERAS programs emphasize early mobilization, optimal nutrition, and patient engagement. In combination with prehabilitation strategies, these programs have been shown to reduce hospital length of stay, complication rates, and overall healthcare costs [24–27].

Despite the demonstrated benefits of conventional rehabilitation programs, access and adherence remain limited due to logistical barriers, mobility limitations, and disparities in healthcare access [17]. Evidence from oncology specific telerehabilitation trials supports its potential with meta-analysis of telehealth exercise-based cancer rehabilitation demonstrated significant improvements in cardiorespiratory fitness and physical activity levels [28]. Similarly, a scoping review of telerehabilitation for patients with cancer reported enhanced functional capacity, cognitive functioning, and QoL (47% of studies), along with reductions in pain and hospital length of stay; it also noted improvements in fatigue, emotional well-being, feasibility, and acceptability [29].

Telerehabilitation, defined as the remote delivery of rehabilitation services using digital technologies, offers a flexible, scalable and patient-centered alternative. Asynchronous telerehabilitation enables patients to engage in tailored exercise and educational interventions at their convenience, without requiring real-time supervision or frequent in-person visits. This model has shown promise in cardiovascular, neurological, and musculoskeletal conditions [30–32] and emerging evidence supports its feasibility in oncology [33].

Preliminary evidence from our group, based on a recent case series of five patients undergoing laparoscopic CRC surgery, showed that an asynchronous multimodal telerehabilitation program integrated into prehabilitation and postoperative phases is feasible, safe, and highly adherent. This exploratory study suggested improvements in functional capacity, strength, body composition, psychosocial factors, and quality of life, with no exercise-related adverse events. Despite its small sample and lack of control group, it offers proof-of-concept data supporting the rationale for the current randomized controlled trial [34].

However, no randomized controlled trials have specifically evaluated the efficacy of an asynchronous multimodal telerehabilitation program, encompassing both prehabilitation and postoperative phases, in patients undergoing CRC surgery. Previous studies have either focussed on synchronous interventions, lacked structured follow-up, or failed to assess psychosocial and adherence-related outcomes [23,35]

Therefore, the aim of this protocol will be evaluated the effectiveness of a structured asynchronous telerehabilitation program, encompassing both prehabilitation and postoperative rehabilitation phases, on physical and psychosocial outcomes in patients undergoing CCR surgery, compared to a traditional leaflet-based control intervention. This protocol seeks to generate novel evidence on digital rehabilitation strategies and their potential to enhance recovery, promote functional independence, and improve long-term outcomes in oncologic surgery patients.

## 2. Methods

### Study Design

This protocol will be designed as a single-blind, parallel-group randomized controlled trial (RCT) with the aim of evaluating the effectiveness of an asynchronous telerehabilitation program integrated into the prehabilitation and postoperative rehabilitation pathway for patients undergoing elective CRC surgery. The study will be conducted according to the Standard Protocol Items: Recommendations for Interventional Trials (SPIRIT) guidelines and will be guided by the Consolidated Standards of Reporting Trials (CONSORT) 2010 recommendations for reporting randomized trials (S1 Appendix).

Participants will be randomly allocated into one of two groups: an intervention group (IG), which will receive an asynchronous multimodal telerehabilitation program, and a control group (CG), which will receive a traditional program through an educational leaflet. The study period will comprise 18 weeks: a 2-week prehabilitation phase, a 4-week postoperative rehabilitation phase, and a single follow-up assessment conducted 12 weeks after completion of the intervention (Fig 1). Participants are currently actively enrolled in the study, with recruitment expected to be completed by the end of 2025. Data collection is anticipated to be completed by April 2026, and the study results are expected to be available by July 2026.

**Fig 1.**
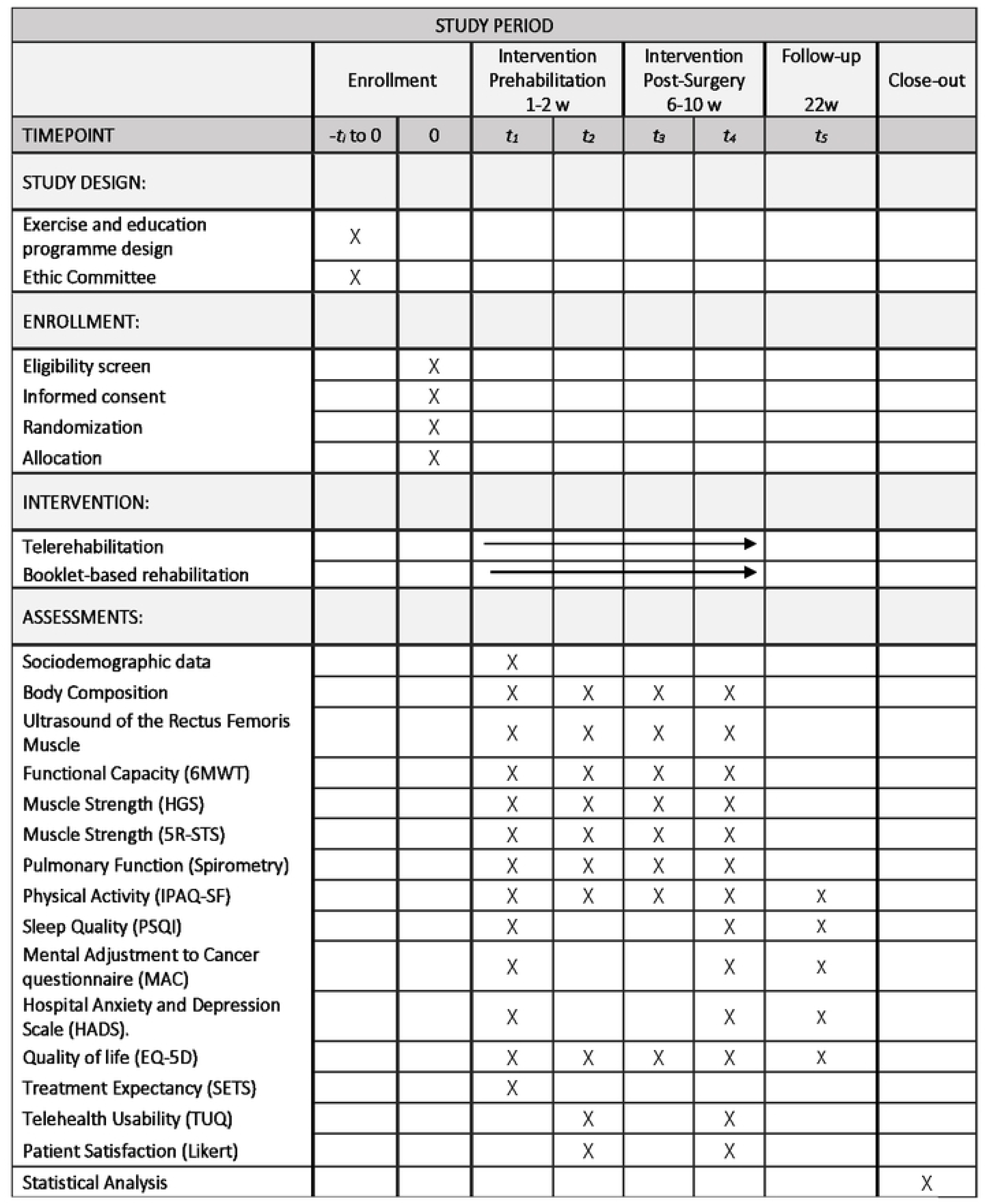
Study Diagram.

This trial protocol has been reviewed and approved by the Ethics Committee of Aragón (Reference: PI23/557) and prospectively registered in ClinicalTrials.gov (NCT06593678.).

### Setting and Population

The study will be conducted in the Department of General and Digestive Surgery of the Royo Villanova Hospital (Zaragoza, Spain), in collaboration with the hospital’s Departments of Endocrinology and Oncology to ensure a multidisciplinary approach. Patient recruitment will be carried out under the direction of the Head of the Department of General and Digestive Surgery, who will identify eligible candidates during preoperative consultations. Participants will be adult patients diagnosed with CRC, scheduled for elective surgical resection, and meeting the predefined eligibility criteria.

#### Eligibility criteria

All potential participants will be informed about the study and must provide written consent before participating. The inclusion criteria will be as follows: 1) Adults aged 18 to 80 years; 2) Patients scheduled for elective CRC surgery at Hospital Royo Villanova; 3) Initial consultation at the Department of General and Digestive Surgery; 4) Functionally independent individuals able to perform walking and pulmonary function tests; 5) Preoperative classification of I, II or III according to the American Society of Anaesthesiologists (ASA) classification and 6) Willingness to participate in the study and signed informed consent.

The exclusion criteria will be: 1) Patients over 80 years old; 2) Preoperative ASA classification IV; 3) Musculoskeletal, inflammatory or other pathological conditions preventing physical exercise; 4) Central and/or peripheral neurological disorders limiting participation in the rehabilitation program; 5) Unstable concomitant cardiac conditions, including cardiac arrhythmias, hypertension, angina or other conditions contraindicating moderate-intensity exercise; 6) Psychiatric disorders diagnosed by a psychiatrist; 7) Lack of access to an internet-enabled mobile device or computer at home; and 8) Refusal to participate or lack of a signed consent form.

#### Allocation and Blinding

Participants will be randomly assigned in a 1:1 ratio to either the IG or CG using computer-generated block randomization (www.randomizer.org), stratified by age and sex. Allocation will be concealed using sequentially numbered, opaque, sealed envelopes handled by an independent researcher not involved in the intervention or outcome assessment.

All participants will be assigned a unique identification code to ensure confidentiality. The outcome assessor will be blinded to the group allocation, and the researcher conducting the intervention will not be involved in any assessment procedures, preserving single-blind conditions.

### Procedure

The recruitment process will be carried out in the Department of General and Digestive Surgery of the Royo Villanova Hospital (Zaragoza, Spain) under the coordination of the Head of Department. During the first preoperative consultation, eligible candidates will receive a comprehensive information sheet detailing the study objectives, procedures, potential risks and benefits, and contact details of the principal investigator for further questions.

Patients who express an interest in participating will be screened by a trained researcher to ensure that they meet the inclusion and exclusion criteria. Once enrolled, participants will sign the informed consent will complete a baseline evaluation (T1: pre-prehabilitation) conducted by a physiotherapist blinded to group allocation. This assessment will involve the administration of standardized questionnaires and physical performance tests. Each session will last approximately 45 minutes and will be performed in the hospital under consistent conditions to ensure the measurement reliability and minimize external variability.

After the initial assessment, participants will be randomized into one of two study groups: (1) the IG, which will follow a structured asynchronous telerehabilitation program; or (2) the CG, which will receive a leaflet with the same rehabilitation protocol.

Participants allocated to the IG will receive access to the HEFORA platform, asynchronous digital tool designed for therapeutic exercise prescription and education. A physiotherapist will support each participant to install and navigate the platform on their personal mobile phone, tablet, or computer, and will provide instructions on how to access the exercise videos, complete session logs, monitor progress, and communicate with the clinical team. Participants of the CG will receive a leaflet containing equivalent educational content, exercise guidelines, and progression instructions for autonomous completion of the rehabilitation program.

A schedule of enrolment, interventions, and assessments is outlined in Fig 2.

**Fig 2.**
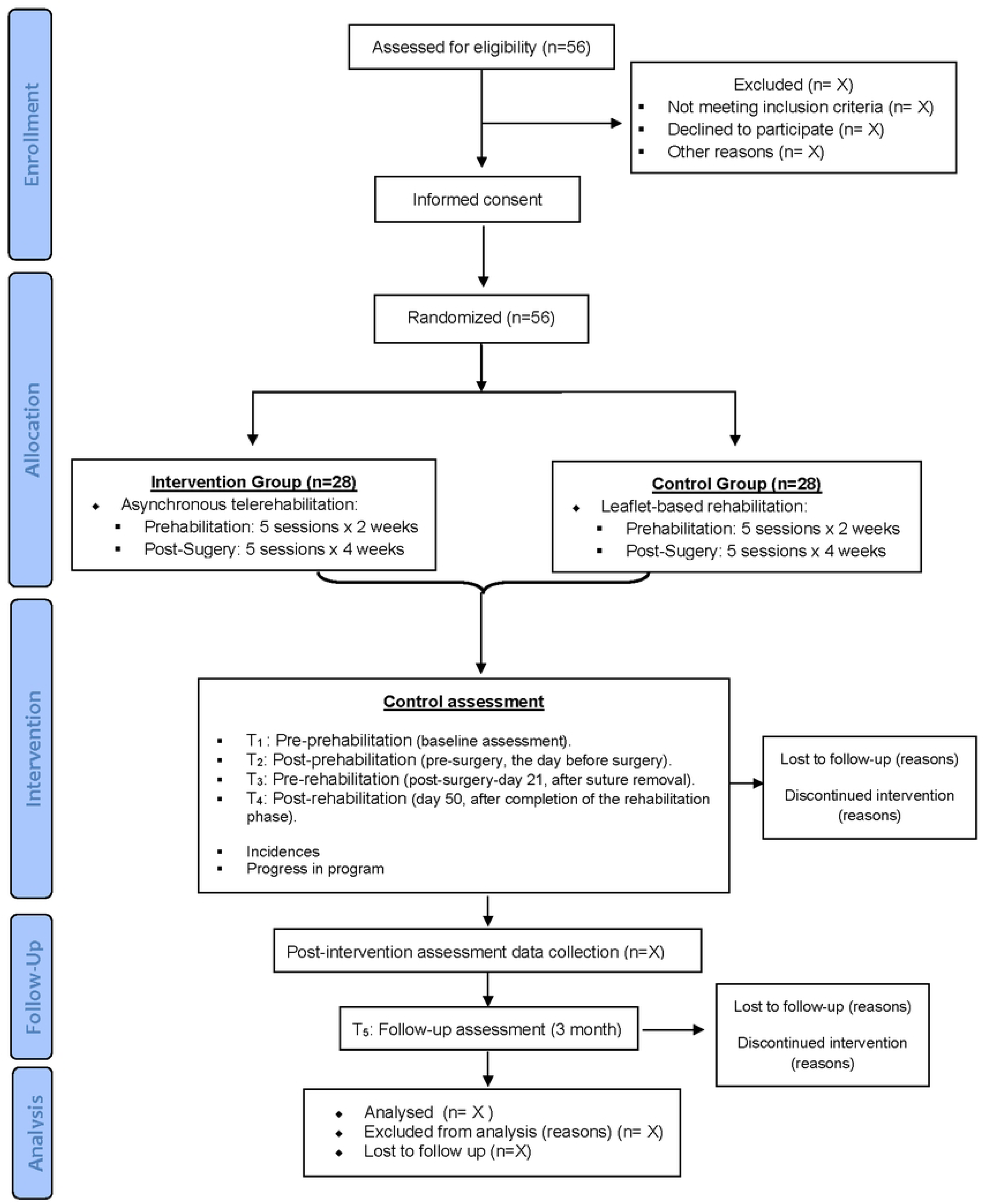
Schedule of enrolment, interventions, and assessments.

All participants will be instructed to maintain a daily exercise log (either digital or paper-based), in which exercise adherence will be documented. This will be performed by completing, a structured adherence diary recording whether each prescribed exercise session was performed fully, partially, with modifications, or not performed, post-session fatigue using a 0–10 numerical rating scale, and any adverse effects. Exercise progress and safety will be monitored asynchronously via the HEFORA platform in the IG, and in the CG through medical review and verification of the adherence diary during the scheduled clinical follow-up visits at the evaluation time points.

Study outcomes will be evaluated at five key time points: Time 1 (T1): pre-prehabilitation (baseline assessment); Time 2 (T2): post-prehabilitation (pre-surgery, the day before surgery); Time 3 (T3): pre-rehabilitation (post-surgery-day 21, after suture removal); Time 4 (T4): post-rehabilitation (day 50, after completion of the rehabilitation phase) and Time 5 (T5): follow-up (three months after T4 by telephone assessment).

Assessments at T1, T2, T3, and T4 will be conducted in person by the same physiotherapist, who will remain blinded to group allocation throughout the study. All evaluations will follow a standardized protocol to ensure consistency across time points. The final follow-up (T5) will be conducted remotely using a structured telephone interview to assess long-term psychosocial well-being and health-related quality of life.

### Intervention

The intervention will consist of a six-week multimodal telerehabilitation program, structured into two phases: a two-week prehabilitation phase and a four-week postoperative rehabilitation phase. The program has been developed in accordance with international clinical guidelines for the perioperative rehabilitation in CRC patients [18,19].

The duration of the prehabilitation period will be limited to two weeks due to the logistical constraints of the public healthcare system, which determine the scheduling of elective CRC surgeries. During this phase, all participants will follow the institutional ERAS protocol [36], This includes preoperative education sessions (about surgical procedures, the expected postoperative course, and self-care strategies), perioperative medical optimization (e.g. medication adjustment and nutritional assessment), and a structured physical training component to improve aerobic capacity, muscular strength, and mobility prior to surgery. In addition, participants will be advised to follow a low-fiber diet in the eight days prior to surgery to reduce intestinal residue, improve intraoperative vision, and reduce the risk of postoperative complications.

Within the ERAS framework, the physiotherapy team will monitor and will deliver the rehabilitation intervention throughout both the prehabilitation and postoperative phases.

#### Intervention Group (IG)

the participants allocated to the IG will access the program through HEFORA, an asynchronous telerehabilitation platform available via mobile application or web interface (HEFORA, Fisio Consultores, Zaragoza, Spain). Each exercise will be presented as a video demonstration with detailed written instructions. The program will be individually tailored according to each participant’s baseline functional assessment and structured into three progression levels (basic, intermediate, advanced) for each exercise category. Initial allocation to a level will be determined by baseline performance, comorbidities, and symptom profile. Progression will be re-evaluated weekly by the assigned physiotherapist, based on participants’ self-reported exertion, symptoms, and adherence data recorded on the HEFORA platform.

Exercise intensity will be guided using a modified Borg Rating of Perceived Exertion scale (target range 3–7) to ensure both safety and effectiveness. Within each progression level, training parameters (including sets, repetitions, tempo, and rest intervals) will be adjusted to promote gradual overload while respecting surgical timelines and recovery phases.

The physiotherapy program will include the following core components:

**(1) Therapeutic education**: participants will receive evidence-based information (via video or printed materials, depending on group allocation) on the principles of prehabilitation, self-care during the postoperative period, strategies for early mobilization, and healthy lifestyle recommendations to promote recovery and autonomy.
**(2) Respiratory exercises**: a daily regimen of breathing exercises will be prescribed, including diaphragmatic breathing, costal expansion techniques, controlled breathing patterns, and incentive spirometry. These exercises aim to improve pulmonary function and reduce the incidence of postoperative pulmonary complications.
**(3) Aerobic exercise**: participants will be encouraged to engage in moderate-to-vigorous aerobic activity for a total of 150 to 300 minutes per week, tailored to their physical condition and surgical timeline, with the goal of enhancing cardiovascular endurance and general fitness.
**(4) Strength training**: resistance exercises targeting major muscle groups will be performed at least four times per week. Typical movements will include bent-over rows, wall push-ups, arm curls, front and lateral shoulder raises with weights, squats (including wall squats), lunges, hip flexor activations, and gluteus maximus and medius strengthening. Training will progress in a structured manner, increasing from 2–3 sets of 8 repetitions to 3 sets of 15 repetitions as tolerated. The program will be updated weekly based on self-reported exertion, symptoms, and recovery status. Adjustments to exercise type, load, or frequency will be made by the physiotherapist responsible for the intervention.

Adherence in the IG will be digitally monitored via the HEFORA platform. Each exercise will include a “Done” button to confirm completion, and participants will be able to record partial completion, reasons for missed sessions, and post-session fatigue using a 0–10 scale. Asynchronous messaging between participants and physiotherapists will enable timely feedback, individualized progression, and the resolution of technical or clinical issues. Automated adherence reminders will be triggered if ≥48 hours elapse without a recorded session. Data from HEFORA will be exported as structured logs to support process evaluation and exploratory analyses.

#### Control Group (CG)

the participants in the CG will follow the same exercise and education protocol as the IG, structured into the same three progression levels (basic, intermediate, advanced). However, all materials will be delivered in the form of a printed leaflet-guide, including step-by-step written instructions and illustrative photographs for each exercise. Training parameters (sets, repetitions, and rest intervals) will be standardized according to the baseline assessment but will not be updated weekly. No real-time feedback, digital monitoring, or asynchronous communication will be provided.

Adherence in the CG will be monitored using a standardized paper exercise diary embedded in the leaflet. Participants will be instructed to complete the prescribed exercises independently at home and to maintain a daily log documenting whether each exercise was performed fully, partially, or not performed, along with perceived exertion and any adverse effects. These diaries will be reviewed by the medical team during scheduled clinical follow-up visits at T2, T3, and T4, and will be verified against the adherence records at each evaluation time point. Any discrepancies between self-reports and clinical observations will be documented to inform sensitivity analyses and to ensure accurate interpretation of adherence.

## Outcome measures

### Primary outcome

The primary outcome of this trial will be the functional capacity, assessed using the Six-Minute Walk Test (6MWT). This submaximal exercise test will be administered following standardized procedures as established by the American Thoracic Society (ATS) and the European Respiratory Society (ERS)[37,38]. The 6MWT quantifies the maximum distance (in meters) that a person can walk in six minutes along a flat, 30-meter corridor, providing an objective measure of submaximal aerobic endurance, cardiorespiratory fitness, and overall physical function [38].

The 6MWT is considered a valid and reliable outcome measure in patients undergoing CRC surgery, as it reflects functional recovery trajectories and physical performance throughout the perioperative period [38,39]. By capturing the integrative physiological response of the cardiovascular, respiratory, and musculoskeletal systems, the test offers a comprehensive indicator of physical capacity and is sensitive to clinically meaningful changes over time [40,41].

### Secondary outcomes

#### Body Composition

Body composition will be assessed using bioelectrical impedance analysis (BIA) with the Tanita BC-601 InnerScan®V (Japan) device. This validated, non-invasive method provides estimates of visceral fat percentage, total body weight, body fat percentage, skeletal muscle mass, and total body water [42,43]. BIA is widely used in both clinical and research settings due to its reproducibility, ease of use and ability to track changes in body composition over time[42,44]. These parameters provide valuable insight into metabolic status, muscle quality and fat distribution, key factors for evaluating the nutritional status and exercise tolerance of patients undergoing CRC treatment.

In addition to BIA, waist and hip circumference will be measured according to a standardized anthropometric protocol to assess central adiposity. These anthropometric indices are clinically relevant as they are strongly associated with metabolic dysfunction, systemic inflammation and cancer-related prognostic factors [45]. In the context of CRC rehabilitation, increased visceral adiposity and low muscle mass (i.e., sarcopenia) have been associated with poorer functional outcomes, higher rates of postoperative complications, and delayed recovery [46,47].

#### Ultrasound Assessment of the Rectus Femoris Muscle

Ultrasound imaging of the rectus femoris muscle will be used to assess skeletal muscle morphology and to monitor changes in muscle mass as a surrogate marker for nutritional status and sarcopenia. This non-invasive, bedside technique has shown high validity and reliability in both oncology and surgical populations and is widely used to detect early muscle wasting in patients undergoing cancer treatment or major surgery [48,49].

The assessments will be performed using a high-frequency (7–12 MHz) linear transducer, with the participant in a relaxed supine position. The transducer will be placed perpendicularly at the lower third (distal third) of the distance between the anterior superior iliac spine (ASIS) and the superior border of the patella on the dominant leg, following standardized anatomical landmarks to ensure reproducibility and accuracy across different time points [50].

The main parameters recorded will include muscle thickness, cross-sectional area (CSA) and echogenicity, the latter being analyzed using grayscale histogram analysis as a marker of muscle quality and intramuscular adiposity. This method allows the monitoring of musculoskeletal adaptations during the perioperative period and the response to therapeutic exercise and nutritional interventions [51].

In patients with CCR, reduced muscle thickness and increased echogenicity of the rectus femoris have been independently associated with higher postoperative complication rates, longer hospital stays, and impaired clinical outcomes [52]. Therefore, serial ultrasound assessments provide valuable complementary data to conventional anthropometric and functional measurements, supporting a comprehensive and dynamic evaluation of the patient status throughout the rehabilitation process.

#### Muscle strength

Muscle strength will be assessed using two validated functional tests: the Handgrip Strength Test (HGS) for upper limb strength, and the Five-Repetition Sit-to-Stand Test (5R-STS) for lower limb function. Both measures are widely used in oncologic rehabilitation as indicators of neuromuscular performance, physical independence, and postoperative recovery potential [53,54]. Upper limb strength will be evaluated using a hydraulic hand-held dynamometer (Jamar® Hand Dynamometer, Sammons Preston Rolyan, Bolingbrook, IL, USA) following standardized testing procedures [55]. The participants will be seated in a shoulder neutral position, the elbow flexed at 90°, and the wrist in a neutral posture. Each participant will perform three maximal voluntary contractions with each hand, holding the grip for three seconds. The highest value (in kilograms) will be recorded for analysis. Handgrip strength is considered a robust surrogate marker for sarcopenia and nutritional status and is predictive of postoperative complications and overall prognosis in CRC patients [56].

Lower limb strength will be assessed using the 5R-STS, in which participants are instructed to stand up and return from a sitting position five times as quickly as possible, with their arms crossed over the chest to avoid upper limb compensation. The total time (in seconds) will be recorded using a standardized chair height (43–47 cm) to ensure consistency. This test reflects the functional mobility and the lower-extremity muscle power, and is strongly associated with fall risk, frailty, and recovery of autonomy after surgery [54,57].

#### Pulmonary Function (Spirometry)

Pulmonary function will be evaluated using standardized spirometry to assess changes in respiratory capacity associated with the telerehabilitation intervention. The two primary parameters to be measured will be: 1) Forced Vital Capacity (FVC): the maximum volume of air forcibly exhaled following a full inhalation and 2) Forced Expiratory Volume in 1 second (FEV₁): the volume of air exhaled during the first second of the forced expiratory maneuver [58].

The spirometry measurements will be conducted using a calibrated digital spirometer, following the technical standards established by the ERS and the ATS [59]. Each participant will perform a minimum of three acceptable and reproducible forced expiratory maneuvers while seated, and the highest consistent value will be retained for analysis. The results will be expressed both in absolute terms (liters) and as a percentage of the predicted values, adjusted for age, gender, height, and body weight[59]. In the perioperative management of CRC, a reduction in lung function is frequently observed, which is associated with a higher rate of postoperative pulmonary complications and delayed mobilization. Specifically, decreased preoperative FVC has been identified as an independent predictor of pulmonary complications such as pneumonia after colorectal cancer surgery [60]. Similarly, a large cohort study in high-risk abdominal surgery patients demonstrated that each 1% increase in preoperative FVC was associated with a 2% reduction in the risk of postoperative pulmonary complications [61]. Spirometry provides a clinically meaningful and objective indicator of respiratory performance and preoperative risk stratification [62].

In this study, spirometry assessments will be performed at several time points to compare the development of respiratory function between the groups. This will help to determine whether participation in the telerehabilitation program, particularly the structured respiratory training component, leads to better maintenance or improvement of lung capacity compared to standard leaflet-based rehabilitation.

#### Physical Activity Level (IPAQ-SF)

Self-reported level of physical activity will be assessed using the International Physical Activity Questionnaire – Short Form (IPAQ-SF). This validated tool is widely used in clinical and epidemiologic research to quantify habitual physical activity in adults and to monitor changes in response to health interventions [63,64].

The IPAQ-SF consists of seven items that measure the frequency and duration (minutes per day and days per week) of physical activity performed in the past seven days across three intensity domains: vigorous activity, moderate activity and walking, and time spent sitting. The answers will be used to calculate total physical activity in minutes per week (Metabolic Equivalent Task, MET-min/week) and allow categorization into low, moderate or high physical activity according to the IPAQ scoring protocol [63].

This questionnaire has shown acceptable reliability and validity for the assessment of physical activity levels in cancer survivors and perioperative patients[65,66]. In the context of CRC rehabilitation, physical activity monitoring is critical to understanding behavioral adaptations, adherence to exercise recommendations, and their potential associations with clinical recovery, functional outcomes, and QoL.

The IPAQ-SF will be administered at each of the five time points (T1–T5), allowing longitudinal analysis of changes in physical activity levels during the intervention and follow-up periods.

#### Sleep Quality (Pittsburgh Sleep Quality Index – PSQI)

Sleep quality will be assessed using the Pittsburgh Sleep Quality Index (PSQI), a widely validated self-report questionnaire to measure sleep disturbances and habitual sleep patterns in the past month [67]. The PSQI is considered the gold standard for assessing sleep in clinical and oncology patients’ populations due to its reliability, multidimensional structure, and sensitivity to change [68,69].

The questionnaire consists of 19 items, which are divided into seven components: Subjective sleep quality, sleep latency, sleep duration, habitual sleep efficiency, sleep disturbances, use of sleep medication, daytime sleep disturbances.

Each component is scored on a scale from 0–3, resulting in a global score ranging from 0 to 21, with higher scores indicating poorer sleep quality. A total PSQI score of >5 is generally used as a cut-off point for clinically significant sleep disturbance [67].

In CRC patients, poor sleep quality has been associated with increased fatigue, mood disturbances, impaired immune function and lower health-related quality of life, particularly during the perioperative and adjuvant treatment phases [70].

Assessment of sleep patterns throughout the rehabilitation process will provide important insights into the psychosomatic impact of telerehabilitation and help to identify potential areas for supportive care interventions. The PSQI will be administered at all five evaluation time points (T1–T5) to monitor changes in sleep quality over time and compare results between groups.

#### Psychosocial factors

Psychological response to cancer and emotional distress will be assessed using two validated instruments: the Mental Adjustment to Cancer questionnaire (MAC) and the Hospital Anxiety and Depression Scale (HADS).

The MAC questionnaire evaluates patients’ cognitive-behavioral coping styles in the context of cancer. It distinguishes between adaptive (positive adjustment) and maladaptive (negative adjustment) responses. Higher scores on the positive dimensions, such as fighting spirit and positive orientation to the illness, reflect active, goal-oriented coping strategies that have been associated with greater psychological resilience and improved treatment adherence [71,72]. Conversely, elevated scores in the negative domains, such as helpless–hopelessness, anxious preoccupation, or fatalism, indicate passive or avoidant coping mechanisms, which are typically associated with emotional distress, reduced QoL, and poorer clinical prognosis [73].

The HADS, a screening tool widely used in oncology for detecting clinically relevant symptoms of anxiety and depression, will be used in the study to assess emotional symptoms. The scale consists of 14 items, equally divided into two subscales: HADS-Anxiety (HADS-A) and HADS-Depression (HADS-D). Each item is rated from 0 to 3, yielding subscale scores ranging from 0 to 21. In clinical practice, the following cut-off thresholds are commonly used: 0–7 (normal), 8–10 (borderline or mild symptoms), and ≥11 (probable clinical case) [74,75].

In CRC populations, elevated HADS scores have been consistently associated with increased psychological burden, diminished QoL, and reduced engagement with treatment regimens. Therefore, the interpretation of MAC and HADS scores will be placed in the context of each patient’s overall clinical profile. In the case of high-risk scores, a comprehensive psychosocial evaluation and referral to mental health or psycho-oncology services when indicated [73].

#### Quality of life EuroQol 5D

Health-related quality of life (HRQoL) will be assessed using the EuroQol-5D (EQ-5D) questionnaire, a validated and widely used self-administered instrument to measure general health status and patient-reported outcomes [76,77]. The EQ-5D captures key domains of physical and psychosocial functioning and is particularly valuable for monitoring changes in health-related quality of life during cancer treatment and rehabilitation [78,79].

The instrument comprises two components: 1) The EQ-5D descriptive system, which evaluates five health dimensions: mobility, self-care, usual activities, pain/discomfort, and anxiety/depression. Each dimension is rated on five levels in the EQ-5D-5L version (ranging from “no problems” to “extreme problems”). The combination of these ratings produces a five-digit health profile, which can be converted into a utility index (EQ-5D index) using a country-specific assessment algorithm. This index ranges from 0.000 (equivalent to death) to 1.000 (full health) and may also include negative values, representing health states perceived as worse than death [77]. 2) The EQ Visual Analogue Scale (EQ-VAS), which captures the participant’s self-perceived overall health status. Participants indicate their current state of health on a vertical scale ranging from 0 (worst imaginable health state) to 100 (best imaginable health state) [76].

In the context of oncological rehabilitation, especially for patients undergoing CRC surgery, the EQ-5D is a widely used tool to assess the treatment burden, the course of recovery and the impact of supportive interventions on physical, emotional and social well-being [78,80,81].

#### Treatment Expectancy (Stanford Expectations of Treatment Scale – SETS)

Treatment expectancy will be assessed using the Stanford Expectations of Treatment Scale (SETS), a validated self-report instrument designed to measure patients’ beliefs and expectations regarding the effectiveness and potential benefits of a therapeutic intervention [82]. This construct has been shown to influence treatment adherence, engagement, and clinical outcomes, particularly in the context of behavioral and rehabilitative interventions [83].

The SETS consists of 6 items, each of which is rated on a 7-point Likert scale ranging from 1 (“strongly disagree”) to 7 (“strongly agree”). The scale captures two dimensions of expectancy:1) Outcome expectancy, the belief that the treatment will be beneficial, and 2) Process expectancy, the belief that the treatment will be delivered competently and professionally [82].

The item scores are summed to produce a global expectancy score, with higher scores indicating more positive treatment expectations. The SETS has demonstrated good psychometric properties, including internal consistency and predictive validity, in populations undergoing various physical and psychological interventions[82,83].

In the context of prehabilitation and postoperative rehabilitation for CCR, patient expectations may play a critical role in motivation, engagement with home-based programs, and perceived benefit of telerehabilitation. By including the SETS, this study will explore the potential moderating effect of expectancy on treatment response [82].

The SETS will be administered at baseline (T1) to capture initial expectations before the intervention and at post-intervention (T4) to investigate possible changes in perception over time.

#### Telehealth Usability (Telehealth Usability Questionnaire – TUQ)

The usability and user experience of the telerehabilitation platform will be assessed using the Telehealth Usability Questionnaire (TUQ), a validated instrument specifically designed to evaluate patients’ perceptions of telehealth systems across multiple domains [84]. The TUQ provides a comprehensive assessment of the practicality, satisfaction and perceived value of remote healthcare delivery tools, and has been validated in a variety of clinical and technological contexts, including applications in cancer rehabilitation and chronic disease management [85,86].

The TUQ consists of 21 items, each of which is rated on a 7-point Likert scale (1 = strongly disagree to 7 = strongly agree), covering six core dimensions: Usefulness, ease of use and learnability, interface quality, interaction quality, reliability, satisfaction and future use [84].

A total and subscale score will be calculated from the item responses, with higher values indicating better usability and user satisfaction. The questionnaire will be administered exclusively to the participants of the IG (telerehabilitation) at the end of the postoperative rehabilitation phase (T4), to evaluate their experience with the HEFORA platform.

Understanding the usability of digital health tools is essential for interpreting adherence and engagement in the telerehabilitation context. The TUQ will provide key insights into the acceptability, accessibility, and perceived value of the asynchronous platform used in this study, which can serve as a basis for future implementation and scalability strategies.

#### Patient Satisfaction

Patient satisfaction with the rehabilitation program will be evaluated using an ad hoc satisfaction survey. This approach has been widely employed in oncology rehabilitation and digital health studies, as patient-reported satisfaction is a key determinant of intervention feasibility, adherence, and long-term sustainability [87,88]. The instrument aims to assess the participants’ subjective perception of the intervention received, whether through the telerehabilitation platform or the standard leaflet-based approach, focusing on the quality of the content, the delivery modality and the overall user experience.

The survey includes two core questions that apply to all participants, which evaluate: 1) Satisfaction with the overall exercise program and 2) Satisfaction with the attention and support received during the intervention.

In addition, participants in the IG will rate: 1) The HEFORA platform (accessibility and ease of use); 2) The exercise video materials; 3) The clarity of exercise instructions and 4) The educational video content. Participants in the CG will provide feedback on the educational leaflet.

Each item will be scored on a 5-point Likert scale ranging from 0 (“very dissatisfied”) to 4 (“very satisfied”). The Satisfaction Survey will be administered at two time points: T2 (post-prehabilitation): To assess early impressions and engagement following the prehabilitation phase, immediately before surgery. T4 (post-rehabilitation): to evaluate overall satisfaction after completion of the full intervention protocol (4 weeks postoperatively).

Measuring patient satisfaction at both stages of the intervention will provide important insights into adherence and participant engagement, factors that are critical to the success and future implementation of telerehabilitation in oncologic surgical pathways [89,90].

##### Sample size calculation

The primary outcome of this trial is the 6MWT distance (meters). The sample size was calculated for a two-arm parallel randomized controlled trial using the standard formula for comparing two independent means [91]. Assumptions included a two-sided α = 0.05, statistical power (1−β) = 0.80, a clinically relevant between-group difference (Δ) of 60 m, based on perioperative colorectal cancer studies reporting minimal clinically important differences of approximately 60–75 m in the 6MWT [22], and a standard deviation (σ) of 80 m, consistent with previous randomized prehabilitation trials in similar populations [92,93]. Under these parameters, the estimated requirement was 23 participants per group (n = 46 in total). To account for an anticipated dropout rate of approximately 20%, the final target sample size was increased to 28 participants per group, resulting in a total of 56 participants. This sample size is expected to provide adequate statistical power to detect clinically meaningful differences in functional capacity while mitigating the impact of potential attrition [94].

##### Data management

A secure and structured database will be developed to systematically track each participant’s progress during the study, including all clinical assessments and outcome measures. Data will be recorded in real time and stored on password-protected, encrypted computers, with access strictly limited to authorized members of the research team directly involved in the study.

All participants will be assigned a unique identification code to ensure the confidentiality and anonymization of personal information. An independent researcher will oversee the data collection process to ensure protocol adherence, monitor data quality and completeness, and safeguard participant safety throughout the study.

Data analysis will only be performed after the completion of participant recruitment has been completed and all outcome assessments have been finalized.

##### Statistical analysis

All statistical analyses will be performed using the Statistical Package for the Social Sciences (SPSS), version 30.0 (IBM Corp., Armonk, NY, USA). A two-sided p-value < 0.05 will be considered statistically significant for all tests.

Descriptive statistics will be used to summarize the baseline characteristics and outcome variables. Categorical variables will be reported as frequencies and percentages, while continuous variables will be expressed as means and standard deviations (or medians and interquartile ranges when appropriate), along with 95% confidence intervals. The Shapiro–Wilk test will be used to assess the normality of continuous variables. To verify baseline comparability between the two groups, parametric or non-parametric tests will be selected based on data distribution: t-tests or Mann–Whitney U for continuous variables, and Chi-square tests for categorical variables.

To evaluate the effect of the intervention over time, general linear models will be applied. If the assumptions of normality and sphericity are met, a repeated-measures analysis of variance (mixed-model ANOVA) will be used to examine time-by-group interactions across the five evaluation time points (T1–T5). When significant main or interaction effects are observed, Bonferroni-adjusted post hoc tests will be used to identify pairwise differences. If the data are non-normally distributed, the Friedman test will be used for within-group comparisons, and significant results will be followed by Wilcoxon signed-rank tests. In these cases, Bonferroni correction will be applied to adjust for multiple comparisons and control for Type I error.

Between-group comparisons at each time point will be conducted using independent-samples t-tests (with Levene’s test for homogeneity of variances) for parametric data, or Mann–Whitney U tests for non-parametric data. Differences in categorical outcomes will be analyzed using the Chi-square test or Fisher’s exact test, as appropriate.

All analyses will be conducted under the intention-to-treat (ITT) principle, including all randomized participants in the groups to which they were originally assigned, regardless of adherence or protocol deviations. Missing data will be handled using appropriate statistical methods, such as last observation carried forward (LOCF) or multiple imputation, depending on the nature and extent of the missingness.

Effect sizes will be calculated to quantify the magnitude of differences observed, using Cohen’s d for parametric comparisons and effect size r for non-parametric comparisons, both for within- and between-group analyses of the main outcomes.

### Ethical aspects and dissemination

This study will be conducted in full compliance with the ethical principles outlined in the Declaration of Helsinki and the guidelines for Good Clinical Practice (GCP). The protocol has been reviewed and approved by the Clinical Research Ethics Committee of Aragón (reference number: PI23/557) and has been prospectively registered in the ClinicalTrials.gov database (identifier: NCT06593678).

Prior to participation, all individuals will receive detailed verbal and written information about the study objectives, the procedures, the potential risks and benefits, and the data protection measures. Written informed consent will be obtained from each participant before enrolment in the study.

To ensure the data confidentiality, each participant will be assigned a unique alphanumeric identification code. All study data will be pseudonymized and stored in encrypted, password-protected digital databases accessible only to authorized study personnel. The final dataset will only be accessible to the statistician responsible for data analysis.

The findings of this study will be disseminated regardless of the direction or significance of the results. Dissemination will include peer-reviewed publications, scientific conference presentations, and open-access repositories, in accordance with the FAIR (Findable, Accessible, Interoperable, Reusable) data principles.

## 3. Discussion

The main objective of this randomized controlled trial will be to evaluate the effectiveness of an asynchronous telerehabilitation program delivered through the HEFORA digital platform, structured in both the prehabilitation and postoperative recovery phases, compared to a standard leaflet-based intervention for improving the physical, psychological and QoL of patients undergoing elective CRC surgery. This proposal responds to the growing clinical need to mitigate perioperative deterioration while overcoming the logistical and geographic barriers that often limit access to center-based rehabilitation programs. By integrating multimodal components into a home-based, digitally guided protocol, this study introduces a patient-centered approach that may improve continuity of care during one of the most vulnerable phases of oncology progression.

The 6MWT has been selected as the primary outcome as it has been shown to be effective in assessing submaximal aerobic performance in surgical and cancer patient populations. Its clinical relevance is underscored by consistent associations between improvements exceeding the minimal clinically important difference (MCID) of 50 meters and a reduction in postoperative morbidity and shorter hospital stays [92,95,96]. By assessing this variable at five perioperative time points, this study builds on the findings of Yang et al. [96], who emphasized the dynamic, non-linear nature of functional trajectories in CRC patients. In addition, a recent meta-analysis supports the efficacy of telehealth-based cancer exercise rehabilitation programs, showing a significant improvement in aerobic performance compared to control groups [28]. These results support the potential of asynchronous telerehabilitation interventions to positively impact the functional outcomes and recovery pathways in CRC patients.

Muscle strength, measured using handgrip dynamometry and the 5R-STS, provides a complementary view of neuromuscular performance and frailty status. Reduced preoperative grip strength is a predictor of complications and delayed recovery [52,97]. Although short-term interventions may not yield large improvements, studies by Minnella et al. and Carli et al. suggest that even modest improvements can influence postoperative exercise capacity when combined with structured rehabilitation [98,99]. Recent evidence from an RCT in breast cancer survivors showed that a 16-week multimodal exercise program, including resistance training, delivered both online and in person significantly improved lean body mass, with no significant differences between delivery formats [100]. These results support the feasibility and potential effectiveness of remote resistance training interventions in oncology care and highlight the relevance of including upper and lower limb strength measurements in the evaluation of the physical impact of telerehabilitation.

Telerehabilitation is expected to positively influence key variables such as body composition, sarcopenia and visceral adiposity, all of which are closely associated with postoperative outcomes in CRC patients[46,101]. Sarcopenia and visceral adiposity have been independently associated with higher postoperative complication rates and lower overall survival in CRC [101,102]. The scientific literature supports that multimodal interventions, particularly those that combine exercise and nutritional support, can mitigate muscle loss and improve patients’ functional status [103,104]. Although Carli et al. [105] and Gillis et al. [106] have shown that such programs can reduce muscle loss, nutritional optimization is often required for sustained benefits, with real-time monitoring of muscle changes emphasizing the need for more sensitive, individualized tools as highlighted by Suárez-Alcázar et al. [107]. In parallel, psychological adaptation, measured with the MAC scale, and emotional distress, assessed with the HADS, will provide information on the psychosocial impact of the multimodal intervention. Maladaptive coping styles such as “helpless–hopelessness” have been associated with poorer recovery, whereas active strategies (e.g. “fighting spirit”) correlate with better treatment adherence and emotional resilience [71,106]. Although physical activity can alleviate symptoms of anxiety and depression [108], evidence from Wu et al. suggests that preoperative anxiety may persist despite functional improvement [109], justifying the inclusion of specific psychosocial support elements in telerehabilitation. However, telerehabilitation has shown promising results in improving psychological adaptation and reducing emotional distress in cancer patients [83,85,110].

Sleep quality plays a central role in recovery from cancer, although it is often underestimated. Using the PSQI allows to examine the relationship between structured exercise and rest-activity patterns [67,69]. In oncology patient groups, poor sleep has been associated with fatigue, immune dysregulation, and impaired QoL [111]. Palesh et al. [112] demonstrated that physical activity interventions can lead to a subjective improvement in sleep, although causality has not yet been established in CRC cohorts. Compared to passive interventions, such as educational leaflets, telerehabilitation provides real-time, interactive, personalized, support that has been associated with greater improvements in patient engagement, symptom management and recovery [85]. This approach is particularly relevant in the context of sleep quality, where structured exercise and psychosocial support, delivered remotely-can positively influence the rest-activity rhythms and reduce fatigue-related sleep disturbances [109].

The assessment of HRQoL using the EQ-5D-5L and the EQ-VAS enables both an objective and a subjective assessment of the global health status. While studies such as those by Dowing et al. [113] and Borchert et al. [114] found that EQ-5D scores often remain high in CRC survivors, self-perceived health status (as reflected in the EQ-VAS) may fluctuate in response to symptoms, pain or mood disturbances. By capturing both domains across five time points, this study aims to provide a more nuanced understanding of recovery [115]. Multimodal prehabilitation and postoperative rehabilitation programs that include physical exercise, nutritional and psychological support have consistently shown better outcomes in surgical oncology, particularly in improving functional capacity, emotional well-being and overall QoL [103,105]. When delivered via telerehabilitation, these interventions maintain their clinical efficacy while improving patient accessibility and adherence, providing a scalable and patient-centered alternative to traditional models [89].

The IPAQ-SF will provide contextual data on patient engagement and behavior change. Perioperative decrements in physical activity are common among patients with colorectal cancer and other surgical oncology populations, but these declines can be mitigated through digital reinforcement strategies [65,116].

Similarly, patientś expectations and perceptions of IG will be assessed using the Stanford Expectations of Treatment Scale, the TUQ, and a customized satisfaction survey. These tools will offer insight into acceptability, usability, and adherence, dimensions that are essential for evaluating real-world scalability. Parmanto et al. [84] and Orlando et al. [117] highlight that usability and perceived benefit are strong predictors of digital adherence, particularly in cancer survivors.

A key limitation of this trial is the brief two-week prehabilitation phase, dictated by the usual interval between diagnosis and surgery in the Spanish healthcare system. While clinically realistic, this short timeframe may limit the physiological gains achievable, especially in muscle strength and functional reserve. Therefore, the findings of this study should be interpreted within the context of this temporal constraint, highlighting the need for future research into flexible or individualized prehabilitation strategies adapted to patients’ baseline functional status and perioperative risk profiles.

To date, no randomized controlled trial has explored asynchronous digital prehabilitation into the postoperative period using a multidimensional assessment strategy that includes physical, psychosocial, behavioral, and experiential outcomes. The present study addresses this gap by providing a structured, scalable model based on validated tools and evidence-based practices [105,118].

If this digitally enabled, patient-centered approach proves effective, it could redefine perioperative care in colorectal oncology, by promoting equitable access, supporting patient autonomy, and addressing both the physical and emotional dimensions of recovery [89,116]. In this way, it could pave the way for wider adoption of asynchronous telerehabilitation as a standard component of value-based cancer care [88].

## 4. Conclusion

This RCT protocol presents an innovative, patient-centered strategy for integrating asynchronous telerehabilitation into the prehabilitation and postoperative recovery of individuals undergoing colorectal cancer surgery. The proposed intervention is based on current evidence and clinical needs and addresses key challenges in perioperative care, such as limited accessibility, lack of time and the need for personalized, multidimensional rehabilitation.

By combining structured therapeutic exercises, patient education and remote monitoring via a digital platform, this study aims to determine whether asynchronous delivery can lead to significant improvements in functional capacity, muscle strength, body composition, psychological adjustment, sleep quality, physical activity and HRQoL.

In contrast to traditional center-based programs, this model emphasizes autonomy, scalability and continuity of care, which may be particularly relevant for patients with logistical or mobility challenges. In addition, the comprehensive five-time point assessment design provides the opportunity to explore the course of recovery in detail and to identify the key phases in which support may be most effective.

If results confirm clinical effectiveness, this intervention could contribute to redefining standard perioperative procedures in oncology by providing an accessible and evidence-based alternative that empowers patients while optimizing outcomes. Ultimately, this protocol is a response to the demand for innovative, equitable and cost-effective rehabilitation strategies within oncology treatment pathways.

## Author Contributions

JM.B.-B., C.J.-S. and S.C. have contributed to the conception and design of this study. JM.B.-B. prepared the original draft of the manuscript. B.C.-P., N.B.-C., L.L.-R, J.L.B.-L, J.A.-S and P.G.-G contributed to the refinement of the methodology and provided critical revisions. C.J.-S, JL.B.-L., and S.C.-C supervised the development of the protocol. All authors participated in the manuscript preparation, including writing, reviewing, and editing, and approved the final version for publication.

## Funding

This research was funded by the Gobierno de Aragón (Number grant: B61_23D).

## Institutional Review Board Statement

The study will be conducted in accordance with the Declaration of Helsinki. The protocol has been reviewed and approved by the Ethics Committee of Aragón (reference number: PI23/557; Acta N° 23/2023).

## Informed Consent Statement

Informed consent will be obtained from all participants prior to inclusion in the study. As this is a study protocol, no individual patient data are reported in this manuscript.

## Data Availability Statement

No datasets have been generated or analyzed yet, as this is a study protocol. Upon completion of the trial, the data will be available from the corresponding author on reasonable request, in accordance with privacy and data protection regulations.

## Conflicts of Interest

The authors declare no conflicts of interest.

## Abbreviations

The following abbreviations are used in this manuscript:

CRC: Colorectal Cancer
WHO: World Health Organization
MDT: Multidisciplinary Team
TME: Total Mesorectal Excision
QoL: Quality of Life
ERAS: Enhanced Recovery After Surgery
RCT: Randomized Controlled Trial
SPIRIT: Standard Protocol Items: Recommendations for Interventional Trials
CONSORT: Consolidated Standards of Reporting Trials
IG: Intervention Group
CG: Control Group
ASA: American Society of Anaesthesiologists
T1: Pre-prehabilitation
T2: Post-prehabilitation
T3: Pre-rehabilitation
T4: Post-rehabilitation
T5: Follow-up
6MWT: Six-Minute Walk Test
ATS: American Thoracic Society
ERS: European Respiratory Society
BIA: Bioelectrical Impedance Analysis
ASIS: Anterior Superior Iliac Spine
CSA: Cross-Sectional Area
HGS: Handgrip Strength Test
5R-STS: Five-Repetition Sit-to-Stand Test
FVC: Forced Vital Capacity
FEV₁: Forced Expiratory Volume in 1 second
IPAQ-SF: International Physical Activity Questionnaire – Short Form
MET: Metabolic Equivalent Task
PSQI: Pittsburgh Sleep Quality Index
MAC: Mental Adjustment to Cancer
HADS: Hospital Anxiety and Depression Scale
HADS-A: Hospital Anxiety and Depression Scale-Anxiety
HADS-D: Hospital Anxiety and Depression Scale-Depression
HRQoL: Health-Related Quality of Life
EQ-5D: EuroQol-5D
EQ-VAS: Visual Analogue Scale
SETS: Stanford Expectations of Treatment Scale
TUQ: Telehealth Usability Questionnaire
ITT: Intention-To-Treat
LOCF: Last Observation Carried Forward
GCP: Good Clinical Practice
FAIR: Findable, Accessible, Interoperable, Reusable
MDC: Minimum Detectable Change

## Notes

### Competing Interest Statement

Dr. Sandra Calvo participated as a researcher in the European project where the HEFORA platform was developed. The other authors declare no competing interests.

### Clinical Trial

ClinicalTrials.gov identifier: NCT06593678

### Funding Statement

Yes

### Author Declarations

Ethics Committee of Aragón (reference number: PI23/557 Acta Nº 23/2023).

